# A note on variable susceptibility, the herd-immunity threshold and modeling of infectious diseases

**DOI:** 10.1101/2021.07.08.21260175

**Authors:** Marcus Carlsson, Jens Wittsten, Cecilia Söderberg-Nauclér

## Abstract

The unfolding of the COVID-19 pandemic has been very difficult to predict using mathematical models for infectious diseases. While it has been demonstrated that variations in susceptibility have a damping effect on key quantities such as the incidence peak, the herd-immunity threshold and the final size of the pandemic, this complex phenomenon is almost impossible to measure or quantify, and it remains unclear how to incorporate it for modeling and prediction.

In this work we show that, from a modeling perspective, variability in susceptibility on an individual level is equivalent with a fraction *θ* of the population having an “artificial” sterilizing immunity. Given that this new parameter *θ* can be estimated, we also derive formulas for *R*_0_, the herd-immunity threshold and the final size of the pandemic. In the particular case of SARS-CoV-2, there is by now undoubtedly variable susceptibility due to waning immunity from both vaccines and previous infections, and our findings may be used to greatly simplify models. If such variations were also present prior to the first wave, as indicated by a number of studies, these findings can help explain why the magnitude of the initial waves of SARS-CoV-2 was relatively low, compared to what one may have expected based on standard models.

## Introduction

Since the fundamental works of Kermack and McKendrick [1–3] compartmental mathematical models (such as SIR, SEIR, etc.) are used to model the spread of infectious diseases. Among other things, these papers introduced the famous *R*_0_-value and showed that, in contrast with human intuition, a very infectious disease will never infect the whole population, but start to decay at a level called the Herd-Immunity Threshold, for which they deduced the famous formula

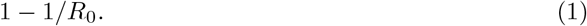

However, prior to the SARS-CoV-2 pandemic, there was no reliable data from a novel virus (affecting humans) on which this prediction could be tested. Unfortunately, this remains largely the case, since e.g. lockdowns and voluntary isolation (which the models can not predict) had a major effect on the spread. Despite this, data from places like Sweden, that did very little to stop community transmission, may indicate that the mathematical models have a tendency to overestimate the magnitude of the wave during a major outbreak.

Several factors are known to have a damping effect on model curves. One such example is variable susceptibility, see e.g. Ch. 1 and 3 in [4], and the articles [5–8]. Similar results have also been established numerically for other heterogeneities, such as age and activity [9]. Curiously, variable infectivity (super-spreaders) do not have any effect on the spread during a major outbreak. In any case, such conclusions are derived using heuristic arguments or by simply testing models, and the mechanisms behind these phenomena remain poorly understood. In particular, since variability in susceptibility is virtually impossible to quantify, it is unclear how to efficiently incorporate it into the models, wherefore predictions of future waves continues to be a major challenge. The same statement also applies to the next pandemic.

Concretely, suppose a novel infectious disease, whose transmission dynamics involves high variability in infectivity and/or susceptibility, is introduced in a well connected network like a large city, and suppose a major outbreak is about to take place. One may then estimate *R*_0_, i.e. a rough estimate of the average number of new infections that one infective gives rise to, from the data series of early cases, using e.g. EpiEstim [10] or [11]. By a contact tracing study one may also estimate the generation time *T*_*generation*_, which is the other parameter needed to run a SIR-model. In such a scenario, one can ask the question if the output of a simple SIR-simulation is a good first order approximation of what is about to come, in the absence of Non-Pharmaceutical Interventions? Is the formula (1) a good indicator of when we may expect the outbreak to start to recede?

Of the prior studies on this topic, the article that comes closest to answering the above questions is Britton et. al. [9], where the authors prove that variations in activity patterns can significantly lower the herd-immunity threshold, in comparison with the *classical* estimate based on (1). An older publication with a similar message is [12]. In both cases, these are purely empirical observations based on models which are intended to illustrate the general effect of population heterogeneity. This effect is not established mathematically and it remains unclear how, and to what extent, different heterogeneities are manifested. In particular it remains unclear how to more accurately predict the herd-immunity threshold.

In the case of SARS-CoV-2, a number of factors such as genetic, cross-reactive immunity and innate immunity, have been shown to provide variation in susceptibility [13–15]. To what extent these factors affected the spread in reality is a complicated question that goes beyond the scope of this paper. However, it is beyond doubt that we presently have great variations in susceptibility due to waning immunity from previous infections (with different variants of concern) along with vaccinations (with various vaccines).

In this work we prove that, as expected, variations in susceptibility have a damping effect on the model curves, whereas variations in infectivity do not (as long as it is uncorrelated with the former, see [6]). More importantly, we also find that the distribution describing *how* susceptibility varies is not needed for accurate modeling; we show that a susceptibility heterogeneous model will behave almost identically to a standard (homogeneous) SIR-model where a portion of the population is totally immune, and that the precise shape of the susceptibility distribution only influences the value of the level of “immunity”. Note that this immunity hence becomes like a mathematical mirage; even if everyone is susceptible to the virus, on a population level it will seem as if a portion of the population have sterilizing immunity. We will refer to such an immunity, needed for accurate mathematical modeling, as “Artificial Sterilizing Immunity” (ASI), and the fraction of the population having it as *θ*. As long as *θ* can be estimated from available data, we show that the actual Herd-Immunity Threshold is indeed lower and given by

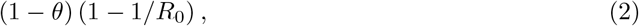

and the final size of the pandemic is also shrunk by the same factor (1−*θ*). We shall also demonstrate numerically that other population heterogeneities, such as those considered by Britton et. al. [9], have an analogous effect, and hence the findings in this paper can be used to significantly reduce the amount of unknowns in a more realistic heterogeneous model for disease spread.

## 1 The mathematics of infectious disease spread dynamics

In order to explain the mathematical findings to a wider audience, we first give an overview of how the basic SIR-model works. SIR stands for Susceptibles, Infectives and Recovered, and is the simplest form of a “compartmental model” used in mathematical epidemiology [16]. In the model, *S, I* and *R* are functions of time *t*, and to illustrate how these are related we shall also introduce the (redundant) function *ν* describing the incidence, i.e. the amount of newly infected each day (not to be confused with *I*, which describes the prevalence). The formula for *ν*(*t*) is at the heart of the algorithm, and in the beginning we simply have *ν*(*t*) = *αI*(*t*), where *α* is a constant that determines how many new cases an average infective gives rise to during a day. If *a* is the average number of daily potentially infectious contacts by an average person, and *p* is the probability that such a contact actually leads to transmission, then *α* = *ap*.

As the amount of susceptibles gradually decreases, we have to modify this by multiplying with the fraction of the population that is still susceptible. If the total population is *N* this fraction is *S*(*t*)*/N* and the formula becomes

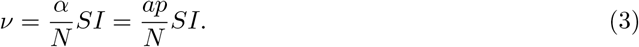

To set up the remaining equations we also need the generation time *T*_*generation*_, i.e. the average time it takes from infection to recovery. The remaining equations are then

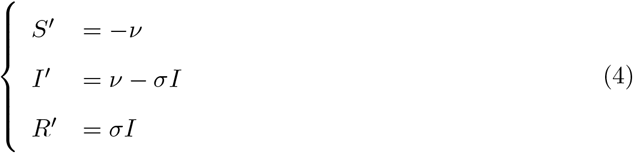

where *σ* = 1*/T*_*generation*_ and ^*′*^ indicates differentiation. The equations are intuitively easy to understand, the incidence continuously gets withdrawn from *S* and added to *I*, and at the same time there is a current of recovering individuals that leave *I* at a rate *σI* and appear in *R* instead.

The SIR-model, and our extensions thereof, are deterministic in the sense that if we run it twice, the output is the same. Such models are believed to work well during major outbreaks, where the law of large numbers applies [4]. All our findings pertain to this situation; for modeling of e.g. the initial phase or household transmission, other types of models are used.

The most natural initial condition for a new disease is to start with *S*(0) = *N* and *R*(0) = 0 (so everybody is initially susceptible and no-one has yet recovered), and set *I*(0) = *n* where *n << N* represents a small number of import cases arriving at time *t* = 0. The value of *n* is completely irrelevant for the shape of the curves that follow, a low value of *n* only gives the equation system a slower start so it takes a while longer for the outbreak to reach a certain value. Once this happens, the curves look exactly the same independent of *n*. See Fig. 1 for some typical examples of *R*-curves and *I*-curves. In this model, *R* is always increasing and levels out on a number which is called “the final size of the pandemic” (see Fig 1a). *S* approximatively looks like *N* − *R*, since the prevalence *I* at any given time is small in comparison with the total population. The incidence *ν* typically looks just like *I*, albeit with a lower magnitude.

**Fig 1:**
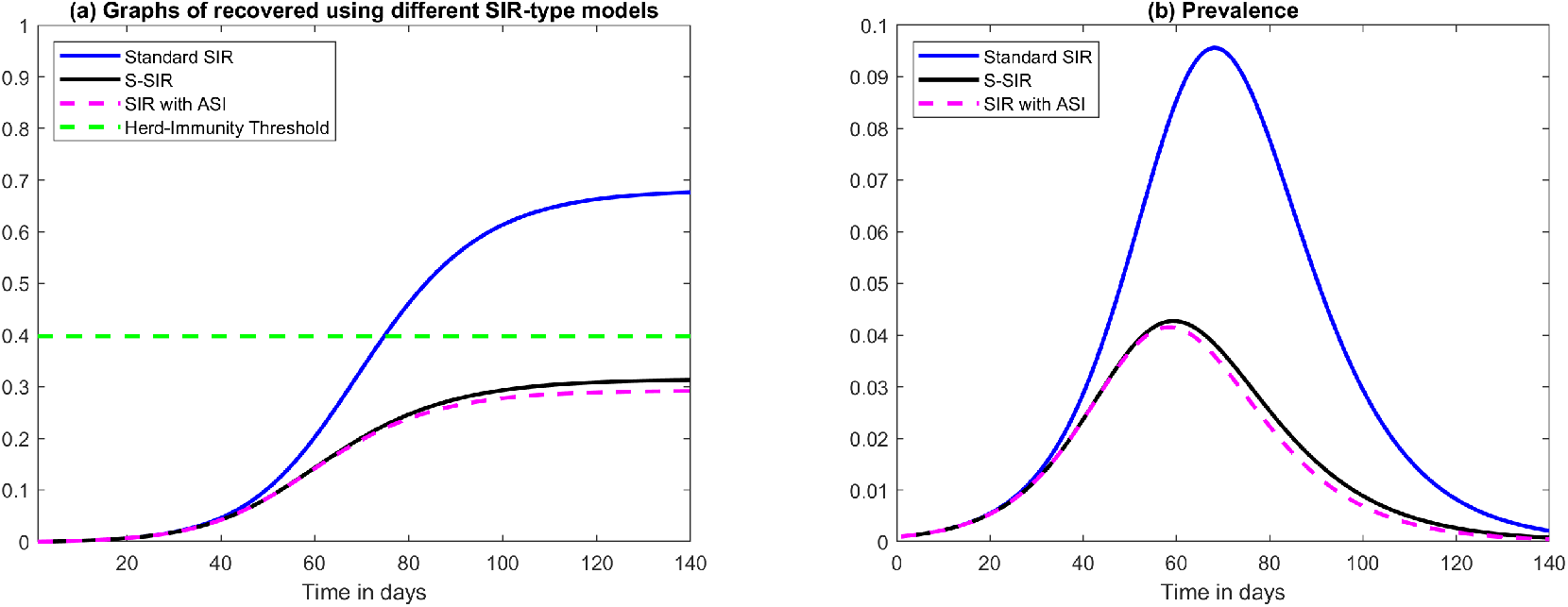
Graphs of recovered *R* and prevalence *I*. (a) Graphs of recovered (as a fraction of the total population) for various SIR-models and a fixed value of *R*_0_ = 1.66. First we display standard SIR, then S-SIR and finally SIR with Artificial Sterilizing Immunity (ASI) with parameters from (8). Note that they start out almost identically but that the latter two bend downwards much earlier than the first, which over-shoots the classical Herd-Immunity Threshold (HIT), whereas the second two stay closely together and level out below the classical HIT. (b) Corresponding curves for prevalence *I*.

### 1.1 Contemporary models for COVID-19

Contemporary models used by professional modeling teams usually contain many more compartments than SIR, for instance relating to age stratification, variable activity levels, geographical regions, compartments for people who need ICU and compartments for people that die. It is common to add a compartment *E* for “Exposed”, incorporating the incubation time. However, as we show in the Section 3, this has a limited effect on the overall prevalence and the final size of the pandemic.

Moreover, compartments relating to severely ill and deaths have a marginal effect, simply because only a small fraction of the infected will end up in these compartments. For example, in Section 3 we show that a simple SIR-model with *R*_0_ = 1.66 and *T*_*generation*_ = 7 peaks at a prevalence of 14%, whereas the corresponding SEIR-model peaks at 13% and the age-activity stratified SEIR by Britton et. al. peaks at 9% [9], i.e. a reduction of about 35% compared to SIR. This is consistent with the findings in [9], where a drop in the Herd-Immunity Threshold of around 30% is observed for the age-activity model, comparing with the prediction (1) based on SIR. Hence, we argue, as a first order approximation to more advanced models, a standard SIR or SEIR-model does a decent job.

For example, the model published by members of the Imperial College COVID-19 response team [17] also has at its root a basic SEIR (see p. 9 as well as Fig. S2 in the supplementary material of [17]), and the same goes for the model [18] used by a renowned Swedish modeling team, which managed to fit the ICU occupancy and deaths with high accuracy during the first wave in Sweden. The latter model also takes into account various regions and interaction patterns between these, but the in-region dynamics is basically a simple SEIR.

The list could be made longer, but the point we wish to make is that, since the output of SIR and SEIR are similar, a standard SIR-model is a good rough approximation to more advanced models, and hence understanding the dynamics of SIR, as we focus on in this paper, is valuable also for analyzing more advanced contemporary models.

### 1.2 Model versus reality mismatch?

Whether or not the more advanced models accurately describe the spread of COVID-19 is hard to determine, since one may always argue that Non-Pharmaceutical Interventions (NPI’s) as well as voluntary behavioral changes have had a major impact. Without claiming to have a definite answer, the case of Sweden is interesting due to its relaxed strategy, which moreover was kept almost constant during 2020-2021. In particular, schools were kept open, people who could not work from home were encouraged to go to work, family members of infected households were obliged to work or go to school, and widespread face-mask use was never implemented, making the country ideal for comparing models with actual data. Due to insufficient testing, the time series of cases is of limited value, but measurements of sero-prevalence from blood samples give valuable information, since it has been established that most people who get COVID-19 also go on to develop anti-bodies [19], and that these antibodies remain for at least 9 months [20, 21]. Results published by the Swedish Public Health Agency [22] indicate that roughly 11% had had COVID-19 in the Stockholm region^1^ after the first wave 2020, which rose to around 22% in February 2021, following the second wave. Also among hospital staff in Sweden (not using face-mask), the prevalence was around 20% [21] after the first wave, in line with observations from infected households elsewhere [23].

However, the model by Sjödin et. al., referred to earlier, predicts a cumulative number of infected people of around 30% after the first wave, despite assuming a 56% decrease in contacts among people of age 0-59 and a 98% reduction among those aged 60-79^2^ (see Fig. 2b, bearing in mind that the Stockholm region has 2.4 million inhabitants). Along the same line, Britton et. al. [9] estimated that the disease could level out at around 43% total infected, in a matter of months. While the authors stress that this is not an actual prediction, it is based on realistic parameters for COVID-19. The famous Report 9 by the Imperial College [24] predicted a total number of 81% infected in a “do-nothing” scenario, based on a more advanced so called “agent based model” that also treats household-contacts separately. According to table 3 in the report, the number of deaths and peak ICU capacity can be reduced by 50% and 81%, respectively, in the most effective NPI-scenario, which certainly goes beyond what was implemented in Sweden. However, as of February 2021, when the original Wuhan-strain was declining [25], these reduced predictions overestimate the actual figure by a factor of roughly 4 (deaths) and 10 (ICU) (when directly translated to Stockholm County).

**Fig 2:**
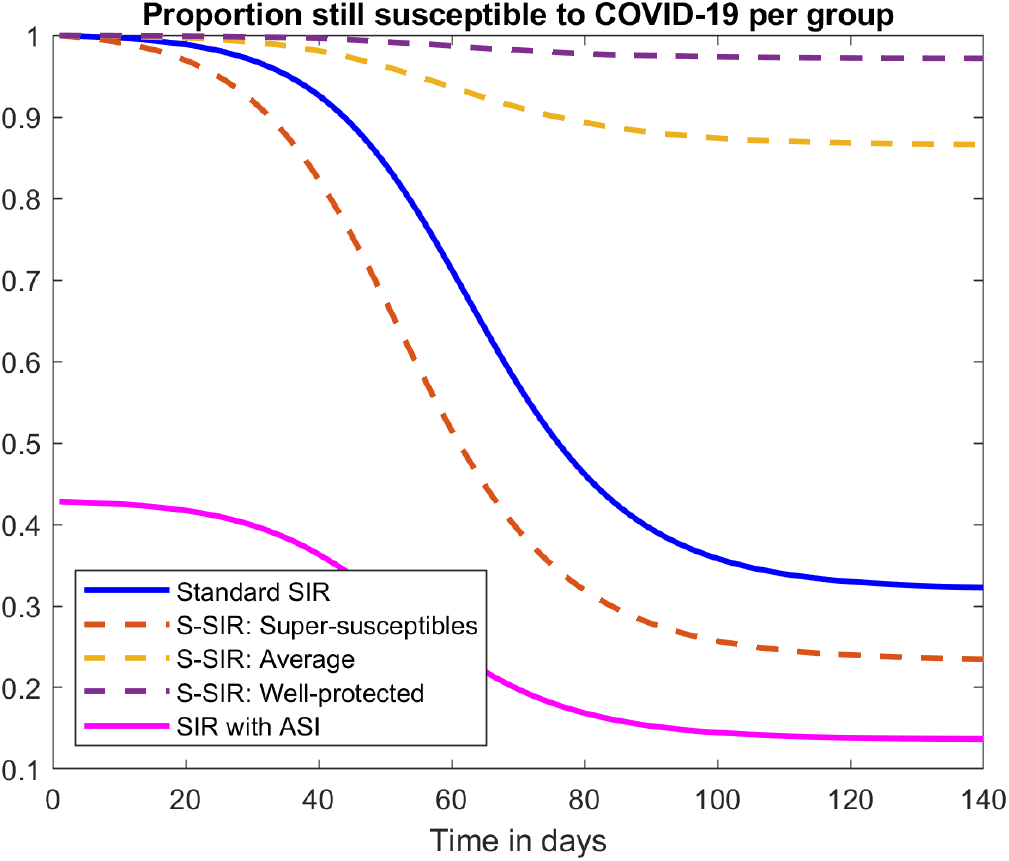
Graphs of susceptibles. *S*−curves corresponding to the 3 graphs in Figure 1. The S-SIR model has three subgroups *S*_1_, *S*_2_, *S*_3_ corresponding to *p*_1_ = 1, *p*_2_ = 0.1 and *p*_3_ = 0.02 (labeled “super-susceptibles”, “normal” and “well-protected”). Note how the spread in the latter two sub-groups level out as soon as it levels out in the super-susceptible group. This rather unexpected behavior is typical. We also remark that the SIR with ASI starts out with 57% immune, which is why the initial value for the pink curve is 43%. This number was chosen using the formula (8).

The point here is not to criticize any particular model, and clearly the case of Sweden can not prove that models are right or wrong, as mentioned initially. However, based on the Swedish numbers compared with the various model outcomes described above, it is a legitimate question whether “contemporary models” have a tendency to overestimate the society spread and final size of the pandemic.

### 1.3 Pre-immunity, super-spreaders and other inhomogeneities

How can we alter the equation system (3)-(4) in order to dampen the curves? The simplest option is to assume that a certain fraction *θ* of the population have some form of sterilizing immunity so that they can not get infected by the virus. Mathematically, this is easily achieved by updating the initial conditions to

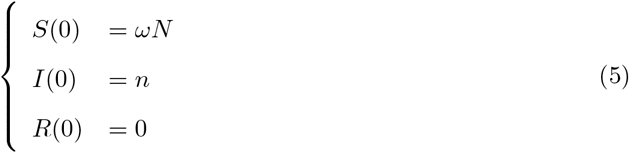

where *ω* = 1 − *θ* is the fraction of initially susceptible. However, this is not very realistic since immunity is usually not binary, i.e. either 100% or 0%. The hypothesis that some people are more susceptible than others is then far more plausible than a binary immunity. In the particular case of SARS-CoV-2, when it was first introduced, the hypothesis that certain individuals had some form of pre-immunity was suggested as an explanation for the, at least for some, unexpectedly mild initial infection waves [26]. This paper lists a number of studies showing that some people had some a priori T-cell immunity. Since then, different articles have demonstrated various mechanisms that make certain individuals more or less susceptible to SARS-CoV-2, e.g. [13–15]. It is also well established that infectivity levels vary dramatically, as mentioned earlier (see e.g. [27]). In addition, this seems uncorrelated to how sick they become; many individuals with very high viral loads are even asymptomatic. In light of this, the most probable assumption is that also the way the virus enters the human is subject to large individual variations.

To make a more realistic model for the spread of COVID-19, or any infectious disease for that matter, it is reasonable to divide the compartments *S* and *I* into a number of subcompartments *S*_1_, …, *S*_*J*_ and *I*_1_, …, *I*_*K*_ where people in each compartment have a different level of susceptibility/infectivity. To see how to set up a corresponding equation system for disease spread, recall that *a* was the amount of daily contacts by one individual. We now let *p*_*jk*_ be the probability that such a contact leads to transmission when an individual in *S*_*j*_ meets one in *I*_*k*_. The incidence *ν*_*j*_ coming from the group *S*_*j*_ then becomes

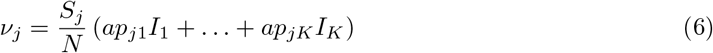

(c.f. (3)). The remaining equations in (4) are easily modified to this new vector setting, see SM Sec. 1 for details. In Section 3 we also discuss other extensions such as SEIR and variable activity levels.

## 2 Main results

The main point of this research is that extensions to both SIR and SEIR of the type mentioned above yield overall curves that are only marginally different from basic SIR, given that a level of Artificial Sterilizing Immunity (ASI) is included. First of all, after setting up the details in Section 1, we prove in SM Proposition 1.1 that the division of *I* into various sub-compartments have no effect whatsoever. In other terms, the existence of “super-spreaders” do not in any notable way affect the dynamics of disease spread. As mentioned initially, this should not come as a surprise to experts, since similar conclusions are scattered in the literature, see e.g. [4, 8, 28]. Removing this layer of complexity, the equations (6) simplify to

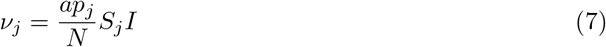

where *p*_*j*_ is the probability of transmission when a susceptible in group *S*_*j*_ encounters an “average” infectious individual. We refer to SM (13)-(15) for the full system of equations, which we label S-SIR for “Susceptibility-Stratified SIR”. It is a very curious fact that the division of *S* into subcompartments can not, in contrast to *I*, mathematically be further reduced to a simpler equation system. However, and this is the key result of this paper, we can prove mathematically that the overall behavior of S-SIR (in terms of prevalence *I* and recovered *R*) differs only marginally from the basic SIR (3)-(4) upon including ASI to the initial conditions, as we did in (5). This is the essence of Theorem 2.1, which is found in SM Section 2. Given probabilities *p*_1_, …, *p*_*J*_, the theorem also provides formulas for suitable values of the transmission coefficient *α* (used to compute the incidence *ν* in (3)) and artificial sterilizing immunity *θ* (used in the initial conditions (5)), as follows:

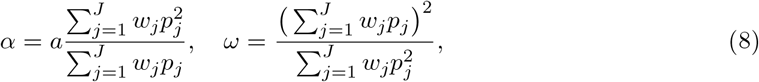

where *ω* = 1 − *θ* and *w*_*j*_ is the fraction of the population initially belonging to *S*_*j*_; *w*_*j*_ = *S*_*j*_(0)*/N*. A simple illustration of these results is found in SM Section 1.3. It is important to be careful with the interpretation of *θ* = 1 − *ω* as a fraction of people who actually have sterilizing immunity, since there is, in reality, not a division of *θN* immune and *ωN* susceptible, which is why we have chosen the acronym ASI; *artificial* sterilizing immunity. These results are illustrated in Figures 1 and 2.

We have observed the same phenomenon also when modeling with SEIR and also when including e.g. different age groups and variable activity levels, following [9]; models with many such layers produce output which seem practically indistinguishable from the output of (3)-(5). We briefly discuss these results in Section 3, which we leave as a numerical observation. In particular, given an observed level of ASI *θ* in a society, it is mathematically impossible to draw any conclusions about how much of *θ* is caused by inhomogeneities in age and behavior, and how much comes from variations in susceptibility.

Incidentally, at the end of each paper [1–3], Kermack and McKendrick stress that a weakness in their model is that they assume uniform susceptibility, which they consider unrealistic in many cases. However, it seems that they never got around to address this issue, and we have not found a rigorous mathematical analysis of how to deal with this situation elsewhere in the literature either. In particular, the formula 1 − 1*/R*_0_ for the Herd-Immunity Threshold (HIT), which stems from their seminal papers, may very well be inaccurate, as demonstrated numerically in [9]. In the coming section we derive a refined version of this formula taking ASI into account.

### 2.1 Formulas for *R*_0_ and the Herd-Immunity Threshold

It is easy to see that the generation time *T*_*generation*_ (introduced below (3)) coincides with the average time an infected individual remains infective. Since *α* is the infection rate, we conclude that *R*_0_ = *αT*_*generation*_ for the standard SIR (3)-(4), assuming a fully susceptible population. However, in the presence of ASI *θ*, the actual infection rate is only (1 − *θ*)*α* and hence the correct formula for the *R*_0_-value

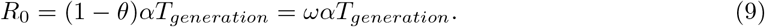

The above value for *R*_0_ is the value that would be estimated by e.g. EpiEstim [10] or [11] from a real time series generated by the model (3)-(4) with initial data (5).^3^ Mathematically, *R*_0_ is defined as the number of new infections that one infected individual gives rise to, before disease induced immunity starts to build up. (To compute this, first solve *I*^*′*^(*t*) = −*σI*(*t*), given *I*(0) = 1, recalling that *σ* = 1*/T*_*generation*_, and then integrate the resulting incidence *ν*, as given by (3), while keeping *S*(*t*) fixed at *S*(0) = *ωN*.) Similarly, one sees that the effective *R*-value, denoted *R*_*e*_(*t*), in the above model is

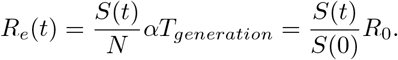

The term “herd-immunity” carry a variety of meanings [29]. In mathematical epidemiology, given a certain model and a novel virus, the Herd-Immunity Threshold is defined as the total number of infective and recovered needed to achieve *R*_*e*_(*t*_0_) = 1. Since *I*^*′*^(*t*) = (*R*_*e*_(*t*) − 1)*σI*(*t*), (recall (4)), we see that this coincides with the point at which the wave of infectious naturally starts to recede. Under such circumstances the disease will naturally die out and, assuming the disease has been temporarily eradicated, any import cases will not spark new outbreaks. For an new outbreak to a known virus, e.g. caused by a mutation or reintroduction in combination with waning immunity, the herd-immunity threshold signifies the proportion that need to get infected during the new wave, in the absence of NPI’s, before the disease start to decline. We denote this value by *H*_*IT*_.

In the SIR-model, it is assumed that individuals mix homogeneously and that recovered individuals have protective antibodies. While it is known that anti-bodies wane over time, at least for SARS-CoV-2, this waning happens much more slowly than the duration of an outbreak [20], and hence this assumption is reasonable for the discussion of the herd-immunity threshold. However, we wish to stress that the waning means that herd-immunity is never a stable condition, but will fade with time.

Assume now that a SIR-model with a certain level of ASI accurately describes a given outbreak. The Herd-Immunity Threshold *H*_*IT*_ then equals *S*(0)*/N* − *S*(*t*_0_)*/N* where the latter is given by solving *R*_*e*_(*t*_0_) = 1 (i.e. the difference between the fraction *S*(*t*_0_)*/N* susceptible at the time *t*_0_ when herd-immunity is reached, and the fraction of susceptibles initially). In the SIR-model with ASI, solving *R*_*e*_(*t*_0_) = 1 yields the equation *S*(*t*_0_)*/N* = 1*/αT*_*generation*_, and so we deduce

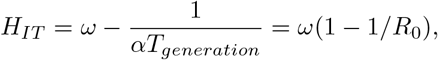

where we used the earlier formula (9) as the definition of *R*_0_. This is the formula for the Herd-Immunity Threshold presented in equation (2) in the introduction. It implies that the classical formula (1), given an estimate of *R*_0_ from e.g. EpiEstim, is over-estimating the Herd-Immunity Threshold. More importantly, it allows to predict *H*_*IT*_, given that the ASI parameter *θ* = 1 − *ω* can be estimated from available data.

That the classical formula may be misleading has been pointed out before [12], and a more recent contribution indicating that the *H*_*IT*_ could be significantly lower than the value (1) is [9]. These works illustrate this by simply testing models that involve heterogeneities (primarily social mixing patterns, not variable susceptibility), and therefore it offers little guidance for actual estimation of *H*_*IT*_. Formula (2) is, to our knowledge, the first time this effect has been mathematically proven and quantified.

To sum up, the results in the previous section imply that a Susceptibility-stratified SIR will behave like SIR with ASI, and the above formula hence applies to this model as well, with *ω* given by (8). It is crucial to note that this applies under the assumption that the immunity is achieved by natural spread. The herd-immunity threshold for vaccinating is still given by the classical formula (1) (assuming the vaccine gives sterilizing immunity). This is shown in SM Section 1.2.

### 2.2 Damping and the final size of the pandemic

As mentioned earlier, several works have established that variable susceptibility have a damping effect on the prevalence. By the above results, this can now can be quantified. Suppose (*S*_0_, *I*_0_, *R*_0_) is a solution to SIR in a homogenous and fully susceptible population (so *S*(0) = *N*), and let *α*_0_ be the corresponding transmission rate. Given a fixed value of ASI *θ*, it is then easy to see that (*S, I, R*) = (*ωS*_0_, *ωI*_0_, *ωR*_0_) is a solution to (3)-(5), where *ω* = 1 − *θ* and *α* = *α*_0_*/ω*. Hence the effect of ASI is really nothing but a rescaling of standard SIR curves. Due to formula (9), the *R*_0_-value equals *α*_0_*T*_*generation*_ in both cases.

It is well known that the final size of the pandemic *π*_0_ = *R*_0_(∞)*/N* in the usual SIR (as well as SEIR) is given by solving 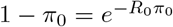 (see [8] and Chapter 3 of [4]). Combining this with the above we see that the final size of the pandemic *π* in SIR with ASI is given by solving

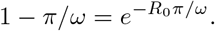

Hence, in combination with our main result about reduction of Susceptibility-Stratified SIR to SIR with ASI, we deduce that the above solution *π* is a good approximation to the final size of the pandemic for S-SIR with *ω* given by (8).

## 3 Extension to more general models

For a disease like COVID-19, with a short incubation period followed by an even shorter infectious period, there is only a marginal difference between modeling using SIR and using SEIR, and hence we believe that the key conclusions of this paper extend to this model as well. Similarly, we have found numerically that more advanced SEIR-models taking variable age and activity levels into account, behave just like SIR if we incorporate ASI. We leave the formal verification of these observations as an open conjecture, and content ourselves with showing some examples.

### 3.1 SEIR

SEIR has two key parameters apart from *R*_0_, namely *T*_*infectious*_ and *T*_*incubation*_, where the former is the average time that a person is infectious and the latter is the time from when a person becomes infected until he or she becomes infectious. Estimates for these vary, we here follow Britton et. al. [9] and set *T*_*incubation*_ = 4 and *T*_*infectious*_ = 3. It then follows that the *generation time* equals

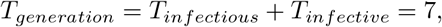

where the generation time is the average time it takes from a person getting infected until that person infects others (see equation (5) in [25]). Note that this is consistent with the choice of *T*_*generation*_ in previous sections.

The reason why SEIR and SIR give almost identical output for COVID-19 is that both are primarily determined by the values of *T*_*generation*_ and *R*_0_. To wit, during a major outbreak, it does not matter if a person is sick for 7 days and infect *R*_0_ people during those 7 days, or if he undergoes incubation for 4 days and then infect *R*_0_ people during the remaining 3 days. As an example, consider Fig. 3 (a); we see a very similar behavior by choosing parameters for SIR and SEIR in accordance with the above formulas (with *R*_0_ fixed). Moreover, by allowing free parameters, SIR can be made to behave almost identically as SEIR (even without involving ASI). Since the exact value for the input parameters are unknown in reality, we argue that it is irrelevant whether one uses SIR or SEIR. Therefore, the observations of this paper should extend to SEIR as well, even if we have not been able to establish this mathematically.

**Fig 3:**
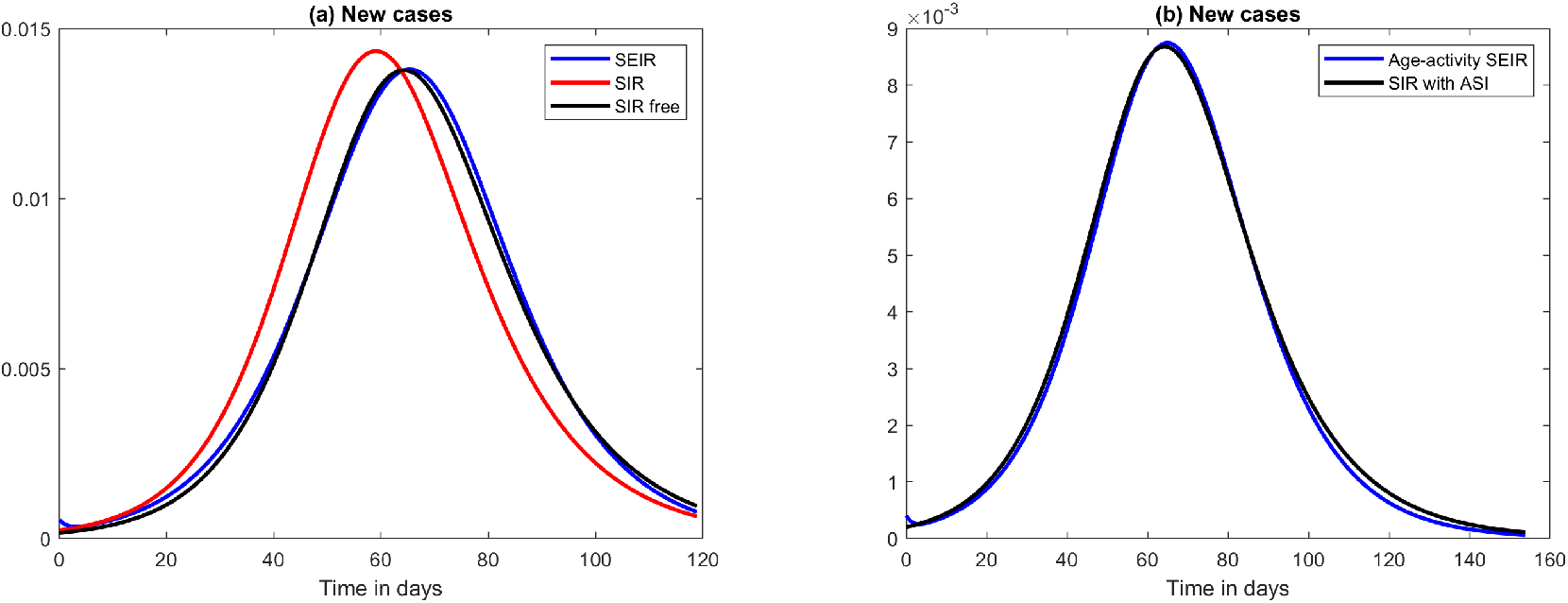
Approximations using SIR with ASI. (a) SEIR with *R*_0_ = 1.66 and *T*_*infectious*_ + *T*_*infective*_ = 7, SIR with the same *R*_0_ and *T*_*generation*_ = 7 and finally SIR with a 1% lower *R*_0_, same *T*_*generation*_ and a lower initial amount of infectives. (b) Age-activity stratified SEIR with *R*_0_ = 1.66 and *T*_*infectious*_ + *T*_*infective*_ = 7 (blue); SIR using the same *T*_*generation*_ but an ASI of 25% and slightly different *R*_0_ (black).

### 3.2 Heterogeneous models

Variable susceptibility is not the only type of population heterogeneity which could manifest itself as ASI on a macro level. In [9] the authors develop a heterogeneous SEIR model taking variable interaction pattern between different age-groups into account, as well as the fact that people in each age-group have varying amount of contacts. We implemented their model and then sought parameters for SIR, allowing for ASI, that would yield a similar output. The result is seen in Figure 3 (b). Again, the difference is so fine that it would be impossible to spot in practice. Henceforth, what may appear as a certain level of population (pre-)immunity in mathematical models may in fact be a mix of various population heterogeneities, in which variable susceptibility is only one ingredient.

## 4 Discussion

There could be many reasons for why certain people are more susceptible than others to infection by a novel virus, ranging from innate and adaptive immunity to cross-reactive immunity from other known viruses as well as genetic differences. For a novel disease, sterilizing pre-immunity, i.e. individuals which are completely immune to without ever having had the virus, most likely does not exist. The key point of this study is that sterilizing individual immunity is not needed in order to observe what looks like sterilizing immunity on a population level, which we have coined ASI; artificial sterilizing immunity. We show mathematically that, in order to have ASI, we only need moderate variation in susceptibility. Moreover, we demonstrate numerically that other types of population heterogeneities, such as variable social mixing patterns, also manifest themselves as ASI. The findings in this paper do not limit themselves to SARS-CoV-2, but basically shows that classical formulas for the herd-immunity threshold and the models for spread of infectious diseases with roots in the famous paper by Kermack and McKendrick [1] are inapt to model any infectious disease subject to large variability in susceptibility, and need to be modified as described in Section 2.1.

The estimation of the herd-immunity threshold *H*_*IT*_ is crucial for efficient disease control management and planning. For example, if a society decides to make a lock-down before *H*_*IT*_ is reached, it is almost certain that the disease will re-emerge, unless strict control of import cases is implemented. The classical formula (1) is still very much in use, despite the fact that it is known to rely on a number of oversimplifying assumptions which may lead to an erroneous indication. We have established a new formula which we prove applies when variable susceptibility is present. Since we show that our simplified model, SIR with ASI, also seems to be a good substitute for models that involve variable social mixing patterns, it is possible that (2) applies more generally than what we are able to prove mathematically.

## Data Availability

Data comes from publicly available data bases. Mathematical code will be posted on GitHub upon acceptance of the manuscript.

## Supporting information

**S1 Supplementary Material**.

## Acknowledgments

We thank Erik Wahlén for fruitful discussions. The research of CSN was supported by the Swedish Medical Research Council (2019-01736).

## Supplementary Material

### 1 Modeling of variable susceptibility and infectivity

We first build a compartmental model taking variable infectivity and susceptibility into account, and then prove that variable infectivity is irrelevant, under the assumption that the two are not correlated (otherwise the statement is false, see e.g. [6]). This is not surprising, see e.g. [8], but it is still worthwhile to prove within our modeling framework. Section 2 is the key contribution where we prove that the resulting susceptibility-stratified model can be further reduced to a basic SIR with ASI.

When modeling variable infectivity and super-spreaders, it is important to distinguish between super-spreaders in the sense that they have an extremely high viral load (possibly combined with lack of symptoms [27, 30]), and *social* super-spreaders, which are persons whose work or behavior puts them in contact with an unusually high number of people. The latter type was discussed in Section 3.2, so we focus here on the former type. We refer to [4] and [31] for the basics of mathematical modeling of infectious diseases.

Assume that the susceptibles can be divided into *J* subgroups, and that the infectives can be divided into *K* subgroups. Let **S** = (*S*_1_, …, *S*_*J*_)^*T*^ be a vector such that 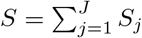 is the total amount of susceptibles, where the symbol *T* denotes transpose, i.e. the operation that turns a row-vector into a column-vector. (All vectors considered here are to be column-vectors for the matrix vector formalism to function.) Similarly **I** = (*I*_1_, …, *I*_*K*_)^*T*^ will represent the various compartments of *I* where 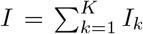. All the above quantities implicitly depend also on time *t* which is a continuous variable. Hence *S*(*t*) and *I*(*t*) denotes the total amount of susceptibles and infectives at a given time *t*, whereas **I**(*t*) and **S**(*t*) contain the amount in each respective compartment. Note that there is no need to introduce various compartments for *R*, since once you recover neither infectivity level nor susceptibility level is important. Let *a* be the daily amount of potentially infectious contacts by an average infective during his period of illness and let *p*_*j,k*_ be the probability that an infectious encounter between a member of *I*_*k*_ and one in *S*_*j*_ leads to transmission, and let *M*_*P*_ denote the corresponding matrix. The rate of new infections in group *S*_*j*_ is then easily seen to be 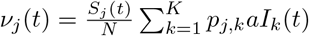, or in vector form

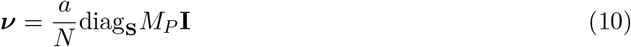

where diag_**S**_ denotes the diagonal matrix with **S**(*t*) and ***ν*** = (*ν*_1_, …, *ν*_*J*_). Since we assume that susceptibility and infectivity are uncorrelated, the total amount of new infections are distributed over the various *I*_*k*_’s in proportion to the fraction of the population that belong to respective group. More precisely, let *x*_*k*_ denote the fraction of the total population that would end up in infectivity group *I*_*k*_, if falling ill, and let **x** = (*x*_1_, …, *x*_*K*_)^*T*^ denote the corresponding vector. The corresponding

SIR-equation system becomes

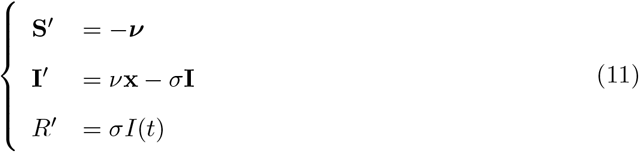

where 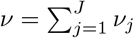 (so it is not a misprint that one is bold and the other not). The initial conditions become

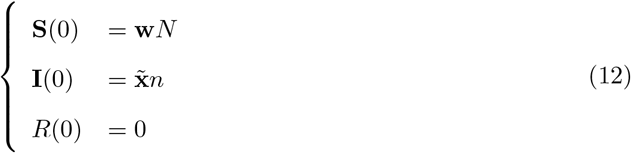

where **w** = (*w*_1_, …, *w*_*J*_)^*T*^ is the fraction of the population in the respective susceptibility group at the onset of the pandemic, *n* is the total number of import cases to the population of *N* individuals, and 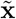 a vector containing the fraction of these in each subgroup of *I*.

These are the equations we propose for disease spread in a model with variable (uncorrelated) susceptibility and infectivity. A weak point of such a model is clearly the difficulty of measuring reasonable values of the transmission probabilities *p*_*j,k*_ in practice. However, this is not needed since the main point of this paper is that the output of any such model is almost identical to the output of the most basic possible SIR if we include ASI. Given a fixed value of *σ* (i.e. the inverse of the generation time *T*_*generation*_), the basic SIR then depends only on two parameters, ASI *θ* and transmission rate *α*. Hence, as long as we believe that there exist numbers such that the above system (10)-(12) would be a good model for accurately forecasting disease spread, we can instead run a basic SIR-model with only two unknown parameters, which are (more) easily fitted to real data. In conclusion, we do not need to know the actual transmission probabilities *p*_*j,k*_, or the contact rate *a*. We now begin to reduce these equation to simpler ones.

#### 1.1 The initial conditions are irrelevant

Even if 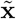 is different from **x**, it will not remain so for long, for the equation for **I**^*′*^ in (11) ensures that at any time point the new cases are distributed according to **x**. Hence it will only take a few generations before the distribution of infected very closely resembles that of **x**. This will happen in the very early stages of the disease progression, and therefore we might as well assume that 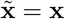 from the beginning.

More rigorously, we can argue as follows; Given the function *ν* describing the total incidence, the solution for *I*_*j*_ is given by

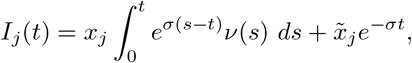

by which it follows that 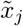 soon becomes irrelevant, due to the exponential decay of the corresponding term 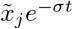.

#### 1.2 Variable infectivity is irrelevant; S-SIR

We now reduce the above equations to the Susceptibility-stratified model (S-SIR) used in the main text. Recall that **I** denotes the vector of all infectivity compartments and *I* denotes their sum.

##### Proposition 1.1.

*Given any solution* (**S, I**, *R*) *to the system* (10)*-*(12) *with* 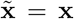, *the triple* (**S**, *I, R*) *solves the system*

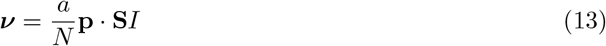

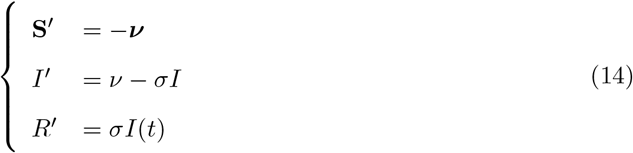

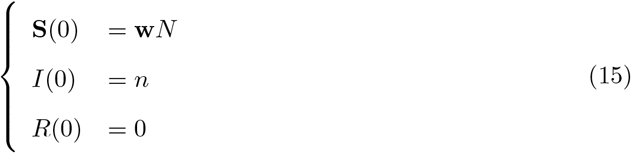

*where* · *denotes componentwise multiplication*, 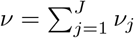 *and* **p** = *M*_*P*_ **x**.

*Proof*. Let (**S, I**, *R*) be given and let ***ν*** be defined by (10). Given a scalar solution to the equation *i*^*′*^ = *ν* − *σi, i*(0) = *n*, it follows that *I*_*j*_(*t*) = *x*_*j*_*i*(*t*) is a solution to the corresponding row in the equation **I**^*′*^ = *ν***x** − *σ***I**. By the uniqueness of such solutions, we conclude that **I**(*t*) = **x***i*(*t*). Hence, since *I* denotes the sum of **I**, we see that *i* = *I*. With **p** = *M*_*P*_ **x** it is easy to see that (10) reduces to (13), and that the equation for **I** in (11) reduces to the middle equation of (14).

Note that the numbers *p*_1_, …, *p*_*J*_ in **p** are between 0 and 1 and can be interpreted as the probability of disease transmission when an “average” infective meets a susceptible in group *S*_1_, …, *S*_*J*_, respectively. We can also compute *R*_0_ given these transmission probabilities via the formula

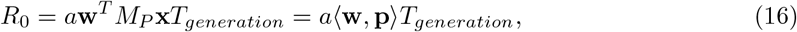

which should be compared with *R*_0_ = *αT*_*generation*_ for the basic SIR (see Section 2.1). The simplest way to derive this is to note that at *t* = 0 we have **S***/N* = **w** so the equation for *I* then reads *I*^*′*^(0) = *a*(**w, p**)*I* − *σI*. This means that an average infective infects *a*(**w, p**) new individuals every day, which in average goes on for *T*_*generation*_ amount of days. Multiplying these two quantities gives (16). The same formula can also be deduced more rigorously using the formal mathematical definition of *R*_0_ as the spectral radius of the next-generation matrix, see e.g. Sec. 5 in [16].

In the S-SIR model (13)-(15), herd-immunity arise when the derivative *I*^*′*^(*t*) becomes negative, i.e. at the point *t*_0_ when the equation 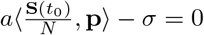. Since it is not possible to theoretically compute the distribution of susceptibles at this point, we can not give a closed formula for the herd-immunity threshold. This underlines the value of the result in the coming section, implying that the value is approximately

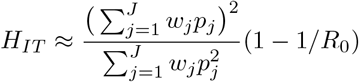

(combine (2) and (8)) The fact that the value is lower than the classical formula (1) is clearly related to the fact that highly susceptible individuals get infected to a greater extent than the other compartments. However, during a vaccination campaign there will be an equal fraction of immunity in in each respective group, i.e. a constant times *ω*. Letting *H*_*IT,vac*_ denote the vaccine induced herd immunity threshold, we thus get the equation

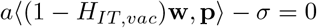

which in combination with (16) implies the classical formula *H*_*IT,vac*_ = 1 − 1*/R*_0_.

#### 1.3 Examples with variable susceptibility

To illustrate, we note that if *J* = 3, then (13) becomes

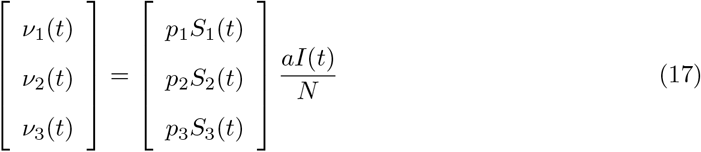

and now the total amount of newly infected becomes

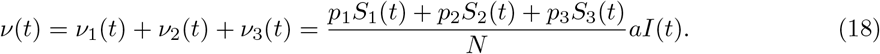

To see why this can not be further reduced to a basic SIR, in a similar way as the proof of Proposition 1.1, note that *ν*_1_ gets withdrawn from *S*_1_, *ν*_2_ gets withdrawn from *S*_2_ and *ν*_3_ gets withdrawn from *S*_3_. If individuals in *S*_1_ are much more susceptible than individuals in *S*_2_, who are much more susceptible than those in *S*_3_ (so *p*_1_ ≫ *p*_2_ ≫ *p*_3_) then the fraction *ν*_1_(*t*)*/S*_1_(*t*) is much larger than *ν*_2_(*t*)*/S*_2_(*t*) and *ν*_3_(*t*)*/S*_3_(*t*), which means that the group of “super-susceptibles” *S*_1_ will become depleted faster. But if *p*_1_ ≫ *p*_2 ≫_ *p*_3_ then the super-susceptibles contribute to the main part of the coefficient in front of *I*(*t*) in (18), and hence it turns out that the disease starts to recede when *S*_1_ becomes sufficiently small, way ahead of what is expected from the standard (homogenous) SIR-model. In other words, while a standard SIR model needs to get above the classical herd-immunity threshold 1 − 1*/R*_0_ before it dies out, the model here can die out as soon as the super-susceptibles are depleted. Thus, once the super-susceptibles are depleted, the disease can die out by itself at fractions much lower than predicted by the classical HIT.

This phenomenon is clearly visible in Figure 2 in the main text. For that experiment, we assumed that *S*_1_ consists of the super-susceptibles, for which *p*_1_ = 1, and further that these constitute 30%, i.e. *w*_1_ = 0.3. Similarly we let *S*_2_ be 60% average group where only one in 10 contacts leads to disease transmission, i.e. *p*_2_ = 0.1, and finally we supposed that the remaining 10% are well-protected with *p*_3_ = 0.02. Clearly, the group of super-suseptibles have a huge attack rate of around 80% (compared with 72% for the whole population in the standard SIR using the same *R*_0_ = 1.66), but as a consequence, the other two groups have attack rates of around 15% and 5%, respectively. This illustrates why traditional estimates for Herd-Immunity Threshold may be overly pessimistic.

To underline that the behavior of S-SIR seen in Figure 1 and 2 is representative, we provide a second example with only 10% super-spreaders and 30% well protected (i.e. we flip the amount in group *S*_1_ with the amount in group *S*_3_). The result is displayed in Supp. Fig. 1. As is plain to see, not much changes; the tiny group of super-susceptibles still “protects” the rest, in the sense that once they are depleted the disease dies out naturally. This group now has a 92% attack rate, as opposed to previously 80% in Fig. 2. Looking at the graph of recovered, we also see that it looks almost as either of the curves in Fig. 1a, only the final size of the pandemic changes to 23%, from 33% for the previous S-SIR and 72% for standard SIR, respectively. Intuitively, it should be possible to find a setting of parameters where the disease burns out among the super-susceptibles, but lives on a bit longer in the “average” group. However, the main finding of this paper is that this intuition is wrong, as we establish in the next section.

**Supp. Fig 1:**
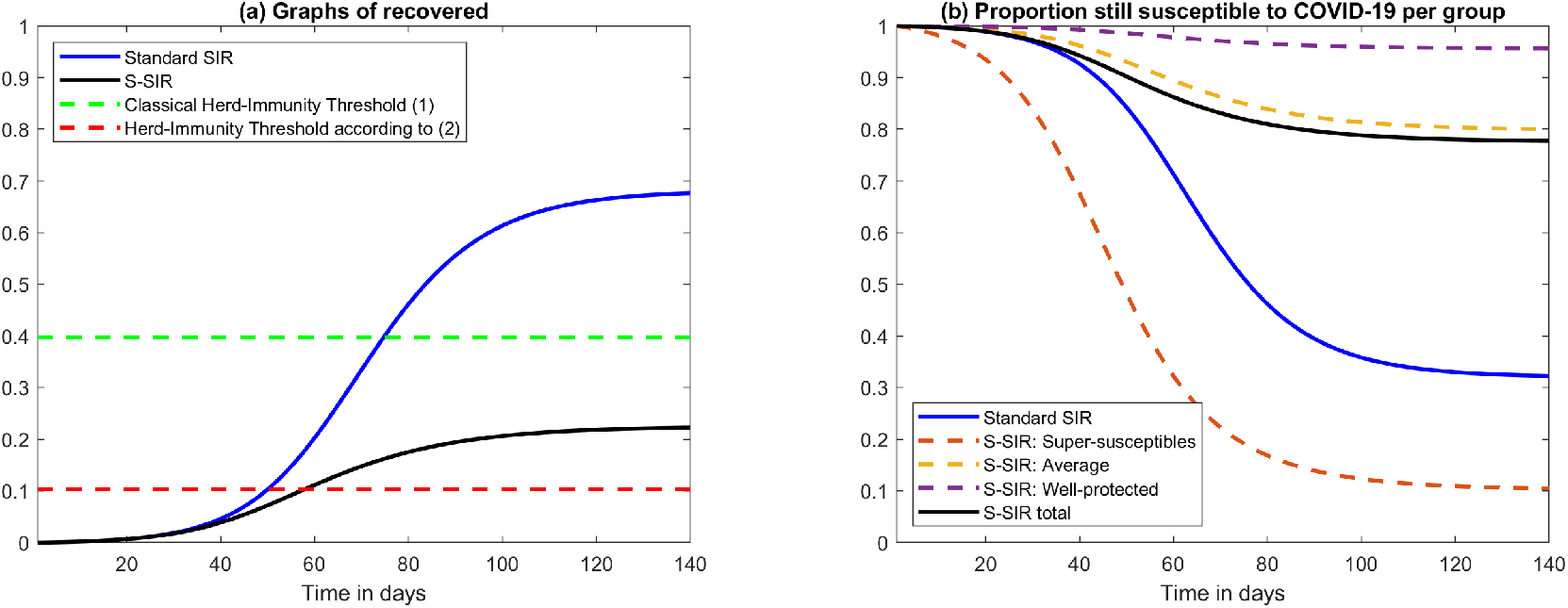
Comparison graphs of *R* and *S* for alternative compartment sizes. (a) Graphs of recovered as fraction of the population for SIR and S-SIR with a fixed value of *R*_0_ = 1.66, along with estimates for the Herd-Immunity Threshold. As expected, S-SIR reaches a final size of the pandemic way below the classical HIT, but about twice that of the HIT as computed according to formula (2) suggested in this paper. (b) Corresponding graphs for susceptibles, which should be compared with Figure 2 in the main text. The model assumes 10% super-spreaders and 30% well protected, as opposed to the situation with 30% super-spreaders and 10% well protected depicted in Figure 2.

### 2 Variable susceptibility is almost equivalent to ASI

We now motivate the claims made in Section 2, in particular formula (8), in the more general setting of the Susceptibility-stratified SIR (13)-(15). We first introduce some notation and explain the intuition behind the formula (8). First consider standard SIR (3)-(5), and let (*S*_0_, *I*_0_, *R*_0_) be a particular solution. We introduce the normalized functions *s*_0_(*t*) = *S*_0_(*t*)*/N, i*_0_(*t*) = *I*_0_(*t*)*/N, r*_0_(*t*) = *R*_0_(*t*)*/N*. These functions solve the system

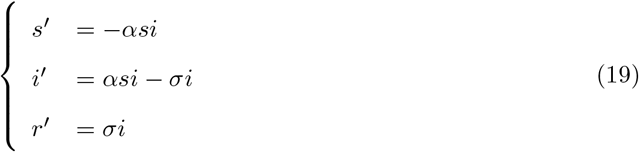

with *s*(0) = *ω, i*(0) = *n/N* =: *ε* and *r*(0) = 0. Note that *s*_0_(*t*) + *i*_0_(*t*) + *r*_0_(*t*) = *ω* + *ε*. Next, we reduce (19) by using the well-known fact that *t* can be replaced by *r* as the independent variable. Let 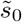 and *ĩ*_0_ be functions defined via 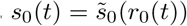) and *i*_0_(*t*) = *ĩ*_0_(*r*_0_(*t*)). By the chain rule the functions 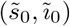 are solutions to the system

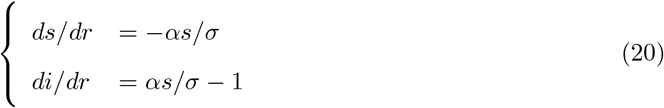

which is easily solved. Since *r*_0_(0) = 0 we have that 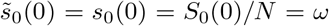, by which we infer that

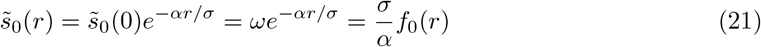

where 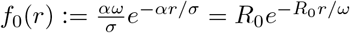. Returning to (20) we see that

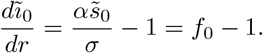

Integrating both sides and using the initial condition *ĩ*_0_(0) = *i*_0_(0) = *ε* gives the solution

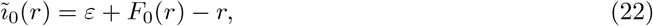

where 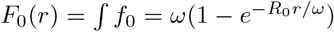. See Supp. Fig. 2 for an illustration.

We now solve the equation for *r*_0_(*t*) in (19), which is separable and can be written

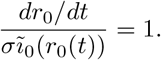

Integrating both sides with respect to *t* and making the change of variables *x* = *r*_0_(*t*) gives

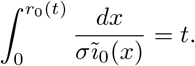

**Supp. Fig 2:**
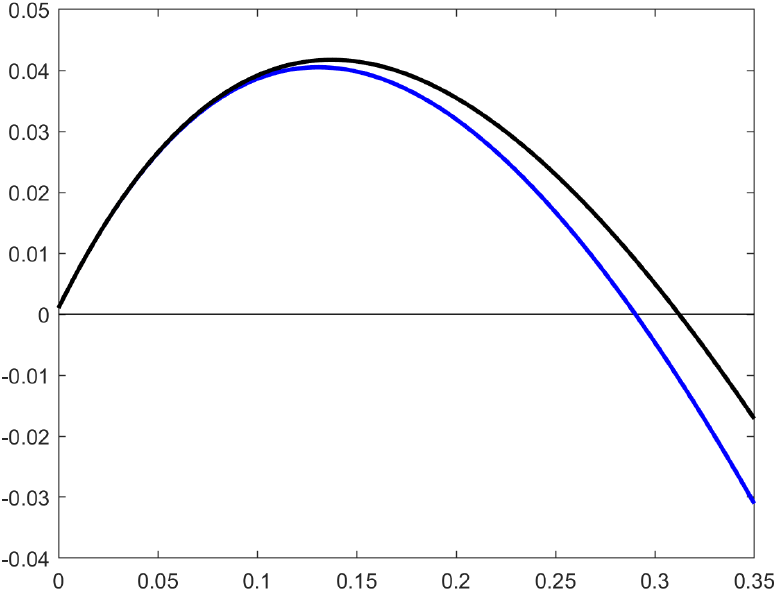
Graphs of *ĩ*_0_ and *ĩ*_1_. The figure shows the solution *ĩ*_1_ (black) for the same example as in Fig 2 and *ĩ*_0_ (blue) using the choice of *α* and *ω* stipulated by (8).

Letting *t*_0_ be the primitive function of 1*/*(*σĩ*_0_) satisfying *t*_0_(0) = 0 we thus obtain the implicit solution *t*_0_(*r*_0_(*t*)) = *t*. Hence, *t*_0_ is also the inverse of *r*_0_ (which exists since 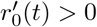 for all *t >* 0), and so

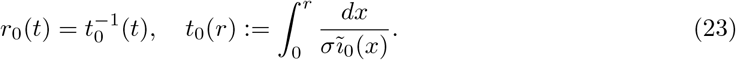

By differentiating (22) we see that *ĩ*_0_ is concave. Let *r*_∞_ be the positive solution of the equation *ĩ*_0_(*r*) = 0, which thus is unique. By standard calculus this gives rise to a non-integrable singularity in (23), and hence 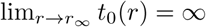. Since *r*_0_ is the inverse of *t*_0_, we see that *r*_∞_ equals the limit lim_*t*→∞_ *r*_0_(*t*), known as the final size of the pandemic. We therefore refer to this number simply as *r*_0_(∞) in what follows. Summing up, *t*_0_ is only defined for values of *r* in [0, *r*_0_(∞)). Note also that as a consequence we also obtain

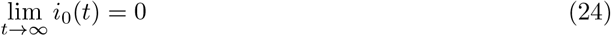

since *i*_0_(*t*) = *ĩ*_0_(*r*_0_(*t*)) so the above limit becomes *ĩ*_0_(*r*_0_(∞)) = 0.

Applying the same arguments to (13)–(15) we obtain first a system in terms of normalized variables

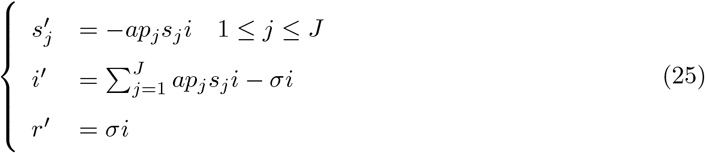

and then an equivalent reduced system of the form

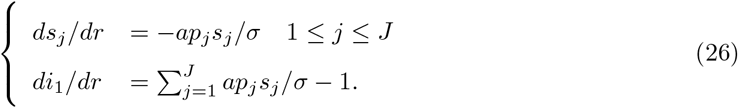

Let 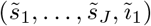 be a solution to (26) with *ĩ*_1_(0) = *ε* and 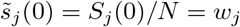 for *j* = 1, …, *J*. Similarly, we will write *r*_1_(*t*) to denote the function described by (23) with *ĩ*_0_ swapped for *ĩ*_1_. As in (21)–(22) we obtain the solutions

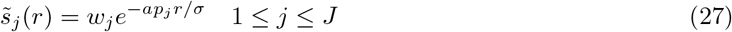

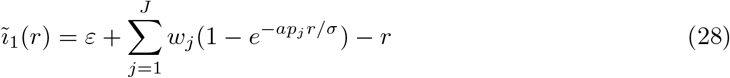

See Supp. Fig. 2 (black curve) for an illustration using the same values as in SM Section 1.3, in particular note that it meets the *x*-axis at the final size of the pandemic *r*_1_(∞), which is seen slightly above 0.3 for the black graph in Figure 1.

The philosophy behind Section 2 and (8), is that we want to pick the parameters *α* and *ω* so that (22) becomes approximately equal to (28). Then, due to (23) we will have *r*_1_(*t*) approximately equal to *r*_0_(*t*) and therefore *i*_1_(*r*_1_(*t*)) approximately equal to *i*_0_(*r*_0_(*t*)). Now, by Taylor’s formula we have

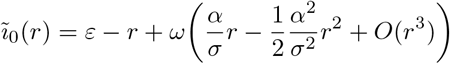

and

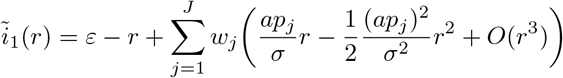

where *O* is the big O notation of Bachman-Landau. Comparing Taylor coefficients we set 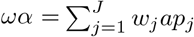 and 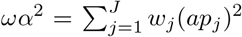 so that the Taylor polynomials of order two of *ĩ*_0_ and *ĩ*_1_ coincide. Solving for *α* and *ω* gives (8), i.e.

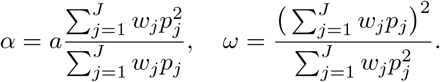

Note here that we always have *ω <* 1. In fact, since 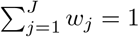 and 0 *< w*_*j*_, *p*_*j*_ ≤ 1 for all *j* it follows that 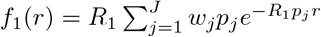, which implies that

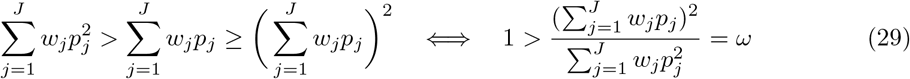

(with strict inequality since at least one *p*_*j*_ is strictly less than one). These formulas can be put in somewhat neater form by introducing *R*_1_ = *a/σ*, i.e. the *R*_0_-value one would have at the beginning of the pandemic if everyone were a super-susceptible. We then get

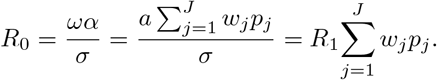

Introducing 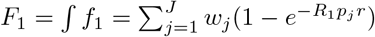 and 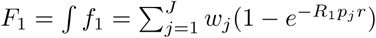, we can write

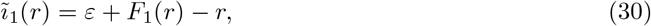

which should be compared with (22). Note that *F*_1_ is engineered to have same first three Taylor coefficients as *F*_0_. Moreover, introducing

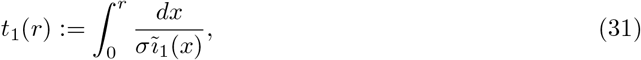

we obtain *r*_1_ as 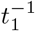 and subsequently 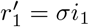.

To summarize this section, we argue that if *F*_0_ ≈ *F*_1_ and *r*_0_ ≈ *r*_1_, then it should follow that *i*_0_ ≈ *i*_1_ since *i*_*j*_(*t*) = *ĩ*_*j*_(*r*_*j*_(*t*)), *j* = 0, 1, and *ĩ*_0_ and *ĩ*_1_ are given by (22) and (30), respectively. To formalize this statement a bit, consider now the more general ODE

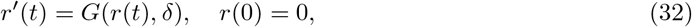

where

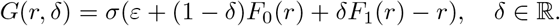

Then *r*_0_(*t*) is the unique solution to (32) for *δ* = 0, while *r*_1_(*t*) is the unique solution to (32) for *δ* = 1. Since *G*(*x, δ*) is independent of *t* and continuous in *δ* for every fixed *x*, it follows that the solution *r*(*t*) = *r*(*t, δ*) of (32) is continuous in *δ*, uniformly in *t* ∈ [0, *T* ] for any *T >* 0, see for example [32] and the references therein. We collect these observations in the following theorem.

#### Theorem 2.1.

*Let r*(*t, δ*) *be the unique solution of* (32). *Then for any T >* 0, *the solution r*(*t, δ*) *is a continuous function of δ, uniformly in t* ∈ [0, *T* ]. *Moreover, we have r*(*t*, 0) = *r*_0_(*t*) *and r*(*t*, 1) = *r*_1_(*t*).

Our numerical simulations have confirmed that indeed, the difference between *i*_0_ and *i*_1_ is small. To illustrate, we divided the population into 100 (equal size) subgroups (as opposed to 3), and assigned probabilities according to the formula 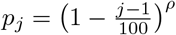 where *ρ* is a parameter to be chosen. This means that we assume a monomial distribution of individual transmission probabilities, and see which exponent *ρ* that seems to give a good fit. In Supp. Fig. 3 we show the curves for *ρ* = 4, 2 and 1 (*p*_*j*_-curve in embedded picture) with respective values of ASI (obtained using (8)) equal to 64%, 45% and 25%. Note that basic SIR with ASI becomes a special case of S-SIR if we choose a function *p* that is either 0 or 1 with a sharp drop at 100*ω*. These curves are also seen in the embedded pictures.

**Supp. Fig 3:**
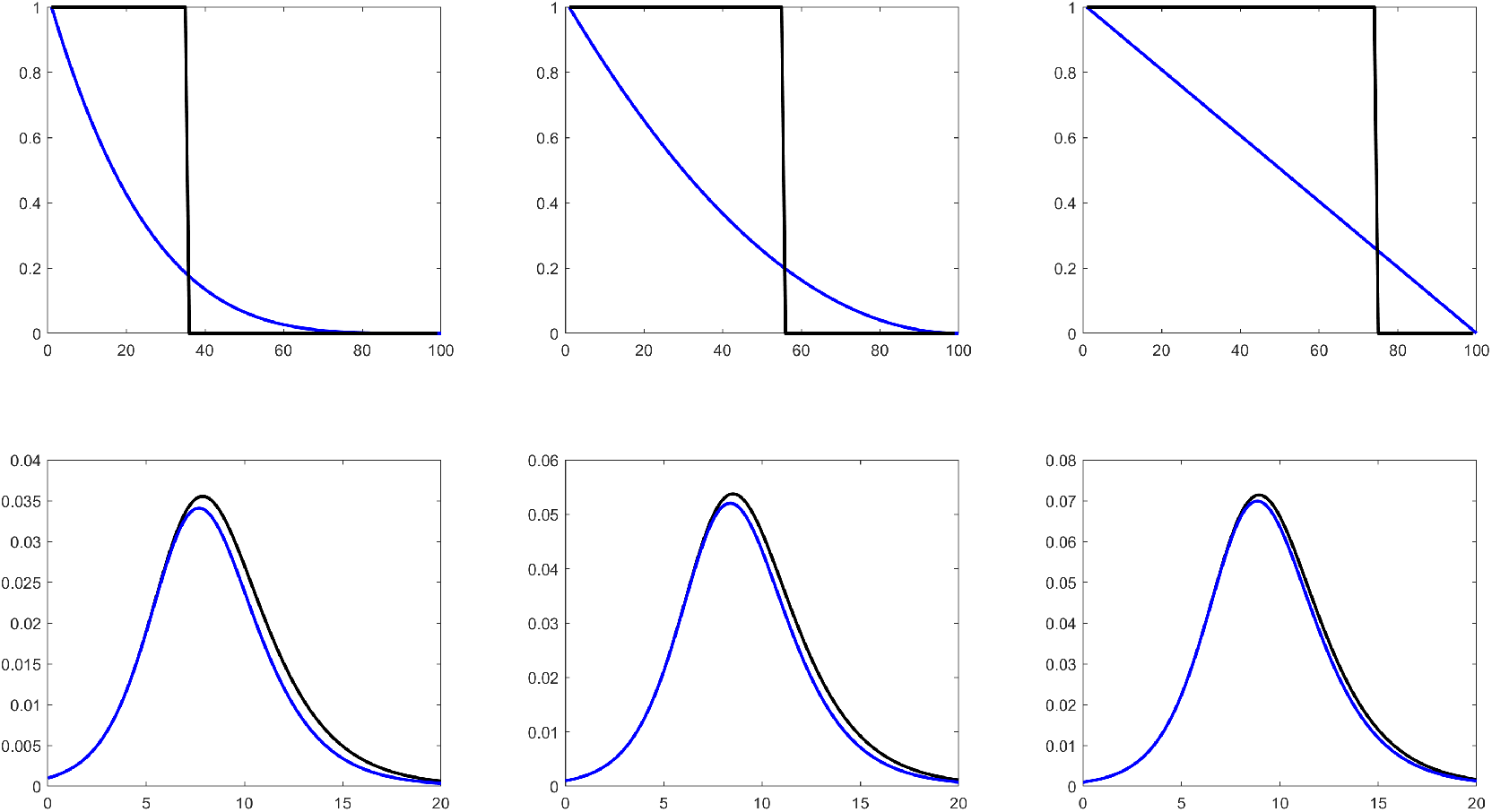
*p*-curves. Top: Various *p*-curves (blue) and corresponding binary immunity curves (black) with ASI obtained from (8). Bottom: corresponding waves of prevalence for S-SIR and SIR with ASI, using parameters obtained from (8).

#### 2.1 Further results and a conjecture

It remains to establish proper estimates proving that the resulting solution (*i*_0_, *r*_0_) is near (*i*_1_, *r*_1_). We first establish the following important inequality, which is the explanation behind that the black curve (from S-SIR) always seems to be a bit higher than the blue one (from SIR).

##### Proposition 2.2.

*It holds that f*_1_ ≥ *f*_0_. *Consequently F*_1_ ≥ *F*_0_, *ĩ*_1_ ≥ *ĩ*_0_ *and dĩ*_1_*/dr* ≥ *dĩ*_0_*/dr. Proof*. The choice of *ω* and *α* is designed so that *f*_1_(0) = *f*_0_(0) and 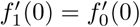. Moreover we have

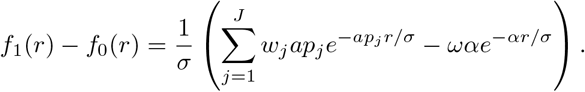

Multiplying with *e*^*αr/σ*^ we obtain 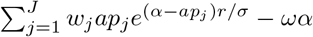 which is a strictly convex function (unless *α* = *ap*_*j*_ for all *j*, which means that all probabilities are the same and the model collapses to the standard SIR model, and there is nothing to prove) that in addition equals zero and has derivative zero at *r* = 0 by construction. By convexity it follows that the function is non-negative, giving *dĩ*_1_*/dr* ≥ *dĩ*_0_*/dr* with strict inequality for *r >* 0. Convexity also implies that the function and its derivative are increasing. The desired inequality follows by integrating *dĩ*_1_*/dr* ≥ *dĩ*_0_*/dr*, keeping in mind that *ĩ*_1_(0) = *ĩ*_0_(0).

##### Proposition 2.3.

*Let r*_0_ *be the solution to* (19) *and r*_1_ *the solution to* (25). *Then r*_1_ ≥ *r*_0_.

*Proof*. Let *g*(*x*) = *σĩ*_0_(*x*). Then 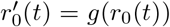 while 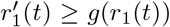 by Proposition 2.2. Since *r*_1_(0) = *r*_0_(0), standard theory of ordinary differential equations then dictates that *r*_1_(*t*) ≥ *r*_0_(*t*) for *t* ≥ 0.

However, we believe there is more to say than what is captured in the above two propositions. Supp. Fig. 4 gives an illustration of how the pairs (*i*_0_(*t*), *r*_0_(*t*)) and (*i*_1_(*t*), *r*_1_(*t*)) evolve with time. Note how the distance seems to be gradually increasing and always points in the north-east direction.

We have not been able to prove the following observation, which in a more concrete way would imply that *i*_1_ and *i*_0_ are close.

##### Conjecture 2.4.

*We have i*_1_ ≥ *i*_0_ *and furthermore*, 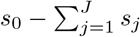 *is an increasing function*.

To understand the implications of the conjecture, recall that the final size of the pandemic solves *ĩ*_*j*_(*r*_*j*_(∞)) = 0, *j* = 1, 2, and that these two points tend to be quite close, see for example Supp. Fig. 2 where the difference *r*_1_(∞)−*r*_0_(∞) is around 0.02 or 2%. Since 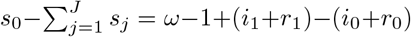 and *r*_1_(∞) − *r*_0_(∞) is the limit of (*i*_1_ + *r*_1_) − (*i*_0_ + *r*_0_) as *t* → ∞, the conjecture implies that both the difference *r*_1_(*t*) − *r*_0_(*t*) as well as *i*_1_(*t*) − *i*_0_(*t*) are bounded by *r*_1_(∞) − *r*_0_(∞).

**Supp. Fig 4:**
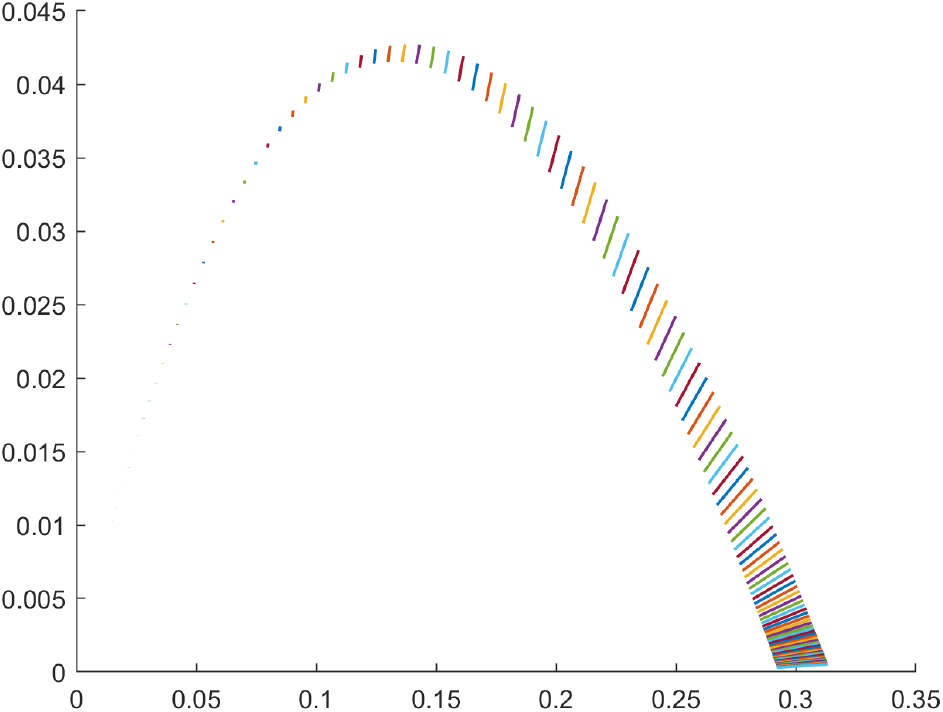
Envelope foliation. Line between pairs (*i*_0_(*t*), *r*_0_(*t*)) and (*i*_1_(*t*), *r*_1_(*t*)) for a grid of *t*-values, using the same parameters as in Fig. 2 for S-SIR and parameters from (8) for SIR. Note that the inner and outer envelope is given by *ĩ*_0_ and *ĩ*_1_ (cf. Fig. 2), and that the slopes are decreasing but positive.

## Footnotes

1 we focus on the capital region since Sweden is a scarcely populated country and geographical constraints are more likely to have a major impact in less populated areas

2 this is for scenario d) which accurately fitted ICU-occupancy and death.

3 up to a minor numerical error caused by the discrepancy between theory and practice

